# Design and Implementation of an End-to-End AI-Driven Colonoscopy Recall Workflow at Scale

**DOI:** 10.1101/2025.07.11.25331400

**Authors:** Aman Mohapatra, Rachel Porth, Si Wong, Heather Hardway, Gail Piatkowski, John Shang, Maelys J. Amat, Sarah Flier, Adam Salsman, Ted Fitzgerald, Ayad Shammout, David Rubins, Amy Miller, Venkat Jegadeesan, Arvind Ravi, Joseph D Feuerstein

## Abstract

**Article Description:** We present a real-world deployment of a large language model-powered colonoscopy recall pipeline that structured over 100,000 patient records during an EHR transition. This end-to-end AI system demonstrated high fidelity, scalability, and a projected prevention of up to 6092 colorectal cancer cases and cost savings between 400 - 670 million dollars.

The rise of structured data elements in Electronic Health Records (EHRs) is a key enabler of improving care quality. However, the transition towards routine use of these fields paradoxically *heightens* patient safety risks due to increased variability in documentation and the use of “placeholder” values pending manual review. For large clinical initiatives such as colon cancer screening and surveillance, misinterpretation of recorded clinical data can be particularly problematic, disrupting risk-adapted recall guidance and potentially exacerbating care gaps. This case study details the development and deployment of a Large Language Model (LLM)-driven workflow to extract and transfer unstructured colonoscopy recall recommendations as part of a larger EHR migration. Utilizing GPT-4 Turbo for the core inference step of a fully integrated pipeline—spanning custom SQL queries, Optical Character Recognition (OCR) of historical PDFs, LLM-based inference, and anomaly detection—we successfully structured and migrated population-wide colonoscopy recall data corresponding to over 100,00 patients and 10 years of clinical care. The pipeline demonstrated high accuracy (Macro F1=1.0 against clinician review), scalability, and cost efficiency. We estimate that use of this workflow—relative to the alternative of a default 10-year reminder from last colonoscopy—may prevent over 6,000 new colorectal cancer cases (a projected cost savings of $400-670 million). Key lessons from implementation include the importance of stakeholder alignment, the necessity of robust quality control at scale, and the technical challenges of expanding optimized LLM inference to a fully-fledged end-to-end clinical workflow.

## INTRODUCTION

Healthcare digitization has driven a shift to structured data capture, enabling precise retrieval, automation, and analytics.^1^ Electronic Health Records (EHRs) increasingly rely on structured fields—such as health maintenance modules and discrete procedural entries—to support quality measures, guideline adherence, and clinical decision support tools.^2^ These developments are foundational for scalable, value-based care delivery.

However, this transformation introduces new operational challenges. Structured data fields often lack seamless continuity with historical unstructured documentation. During EHR transitions or module rollouts, existing free-text clinical data may not be retrospectively structured at scale, leaving critical information fragmented or inaccessible. This creates a transitional period of clinical risk: clinicians assume structured fields are accurate, but historical data may be missing or misrepresented. Prior to our institution’s EHR transition, we predicted thousands of such inaccuracies would be applied to patients undergoing colonoscopy surveillance, highlighting a common problem with broader relevance to other healthcare systems undergoing digitization.^3^ When structured health maintenance fields are relied upon to track surveillance intervals, errors in data migration or field population can result in inappropriate surveillance—either missed recalls or unnecessary procedures.

To accurately facilitate data migration during our EHR transition, we developed an end-to-end large language model (LLM)-powered system to extract, validate, and populate colonoscopy recall fields using historical procedural documentation. Our pipeline identified clinically appropriate recall intervals from unstructured reports, flagged errors and edge cases, and accurately transferred colonoscopy recall data to the new EHR (Fig.1). We demonstrate how this tool fills the fidelity gap created during structured data transitions—bridging the divide between historical workflows and the structured data future.

**Figure 1.**
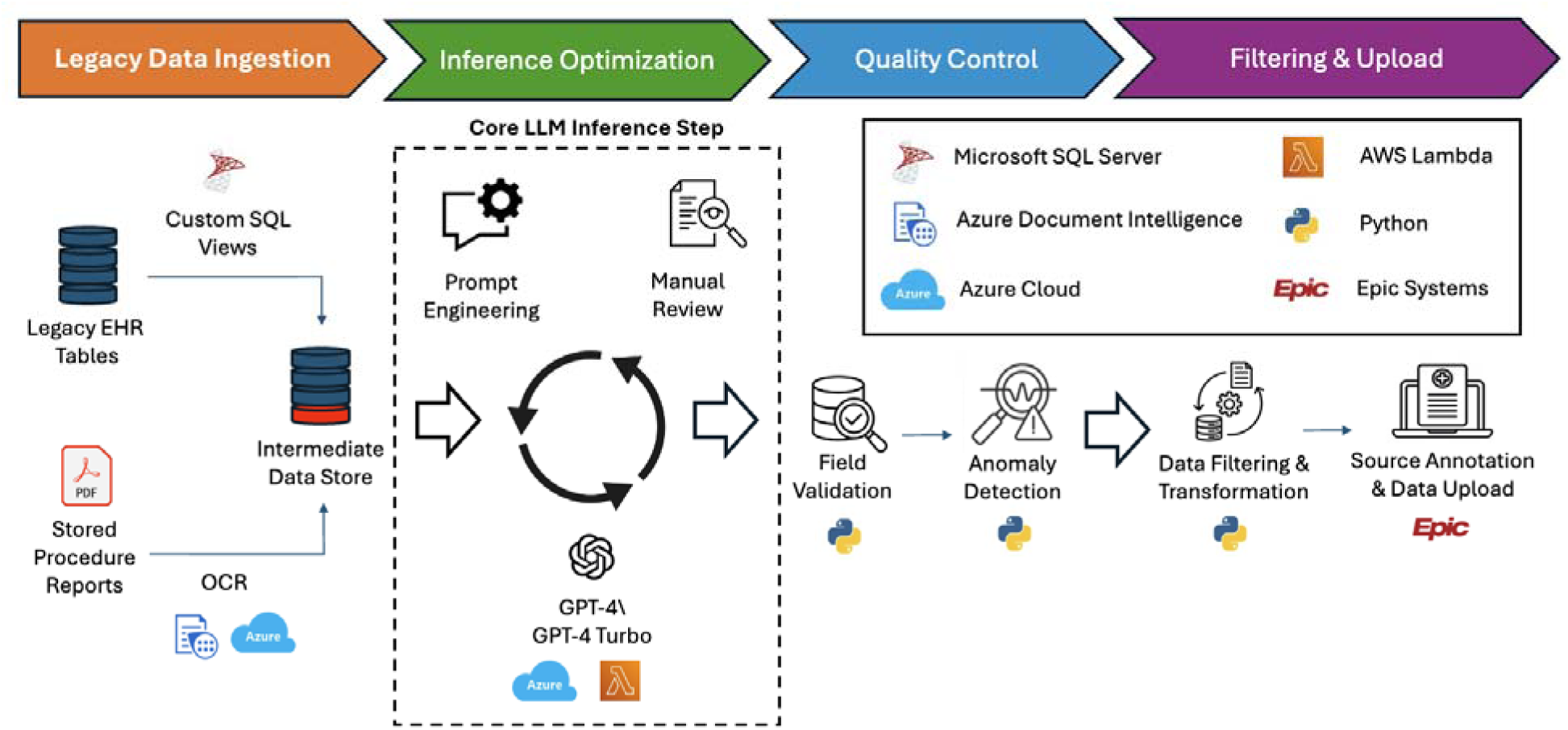
Overview of end-to-end LLM-driven colonoscopy recall pipeline. The recall pipeline development process involved 4 stages, namely 1) legacy data ingestion, in which primary legacy EHR tables were extracted via custom SQL views and merged with stored procedure reports which were separately processed via an OCR pipeline involving Azure Document Intelligence; 2) inference optimization, which is the traditionally emphasized core LLM inference step involving iterations of prompt design, inference, and manual review; 3) quality control, using a combination of field validation and anomaly detection to identify and process inference results at scale; and 4) filtering and upload, in which inference results were transformed into an upload ready format with the addition of a comment citing the source clinical document (i.e., provenance annotation) to aid physician review. EHR = Electronic Health Record, SQL = Structured Query Language, OCR = Optical Character Recognition, AWS = Amazon Web Services. Created by authors using Microsoft Powerpoint with icons sourced from royalty-free image repositories (e.g., Google Images/Flaticon). No copyrighted content was used.

## METHODS

### Project Initiation and Approvals

Key stakeholders—including leadership from hospital administration, information systems, gastroenterology, and quality and safety teams—collaboratively designed an optimal AI-driven recall workflow to transition from our legacy EHR (webOMR) to Epic.

### Preprocessing of Legacy Data

Custom SQL queries generated views from our legacy enterprise data warehouse for patient demographics, endoscopy scheduling, and clinical notes (2014 onwards). A cron job was designed to copy historical endoscopy PDF reports (gGastro) to a project-specific directory. Procedure reports underwent OCR (Azure AI Document Intelligence) to convert PDFs to text.

### Optimization of LLM-Based Recall Inference

We used AWS lambda to test prompts for extracting recall intervals from free text inputs using GPT-4 (gpt-4-0314) and GPT-4 Turbo (gpt-4-1106-preview). Models returned structured JSON recall intervals, including lower/upper bounds (e.g., “3–5 years”) and units for both standard and accelerated intervals(Fig. 2).

**Figure 2.**
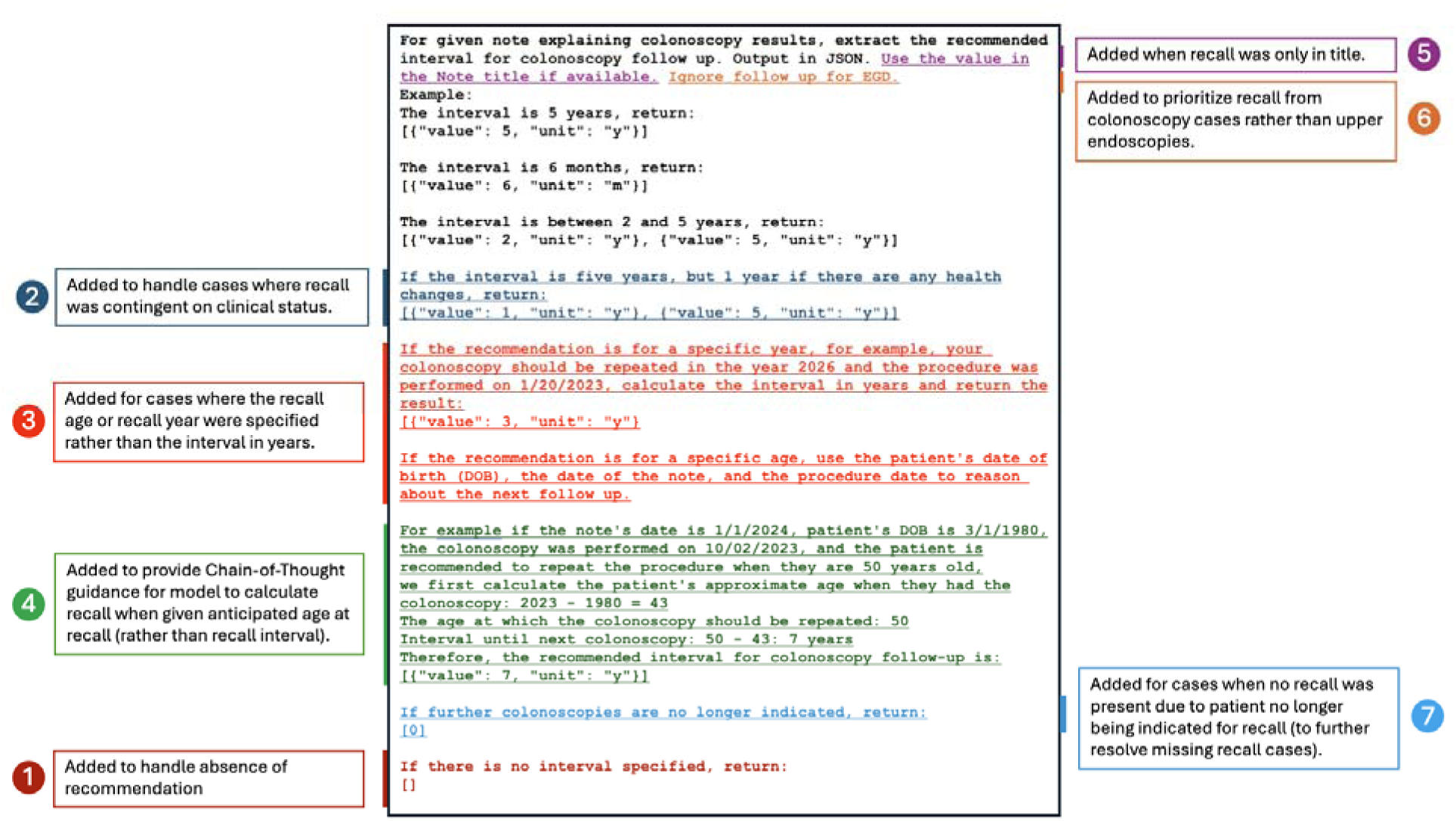
Evolution of a prompt: annotated stages of prompt development for colonoscopy recall inference. The iterative evolution of the prompt inference is shown here, with the base prompt shown in black, and subsequent additions shown in distinct colors along with underlining for emphasis. Clinical context/motivation for each update is shown in the annotations in the margins. Initial QC (N=100 patients) was assessed with prompt revision 2, with subsequent additions following later rounds of QC. Original figure created by authors using data from hospital database (refer to methods section).Visualization created in Microsoft PowerPoint.

When available, recall from patient letters was prioritized as these typically integrate both endoscopy and pathology findings. F1 scores were calculated against manual clinician review.

### Quality Control and Anomaly Detection

We employed three complementary approaches for quality control:

1. **Manual review**: For initial prompt engineering and small-scale assessment (N≤100), we assessed discordance from clinician judgment.
2. **Field validation**: For intermediate and large-scale data, we summarized unique outputs for each component and assessed for invalid results and outliers.
3. **Anomaly detection**: For large-scale review, we derived additional features from primary inputs/outputs (e.g., “age at last colonoscopy,” “age at recall”) and assessed for outliers.

### Postprocessing and Data Upload

Based on quality control assessments, we performed final filtering using custom Python scripts. For transparency, we included an automatically generated comment indicating the source and date of recommendations. The recall list was saved in CSV format for bulk import (Chronicles Import Utilities).

### Workflow Automation

The pipeline was established as a batch process run in multiple phases: 1) first for testing and development, 2) then at population scale two months prior to go-live, and 3) just before go-live to capture interval colonoscopy results.

## RESULTS

### Inference Optimization and Model Selection

As a first step in optimizing our core LLM-based inference step, we began by experimenting with simple prompts to extract structured recall data from procedure reports and patient letters. Because recalls are sometimes specified as ranges, we attempted to extract data as a lower and upper bound (rather than a single value), and with fields for units (e.g., months, years) to accommodate short recall periods. These and future adjustments to the prompt text (e.g., note title parsing, etc.) are shown in Figure 2.

To formally assess the performance of GPT-4 using our updated prompt, we performed inference on procedure reports and patient letters from an unseen sample of 100 patients. The model showed excellent performance on this initial assessment (Fig. 2), with perfect concordance relative to manual clinician annotations (Macro F1 = 1.0; Fig. 3A-B).

**Figure 3.**
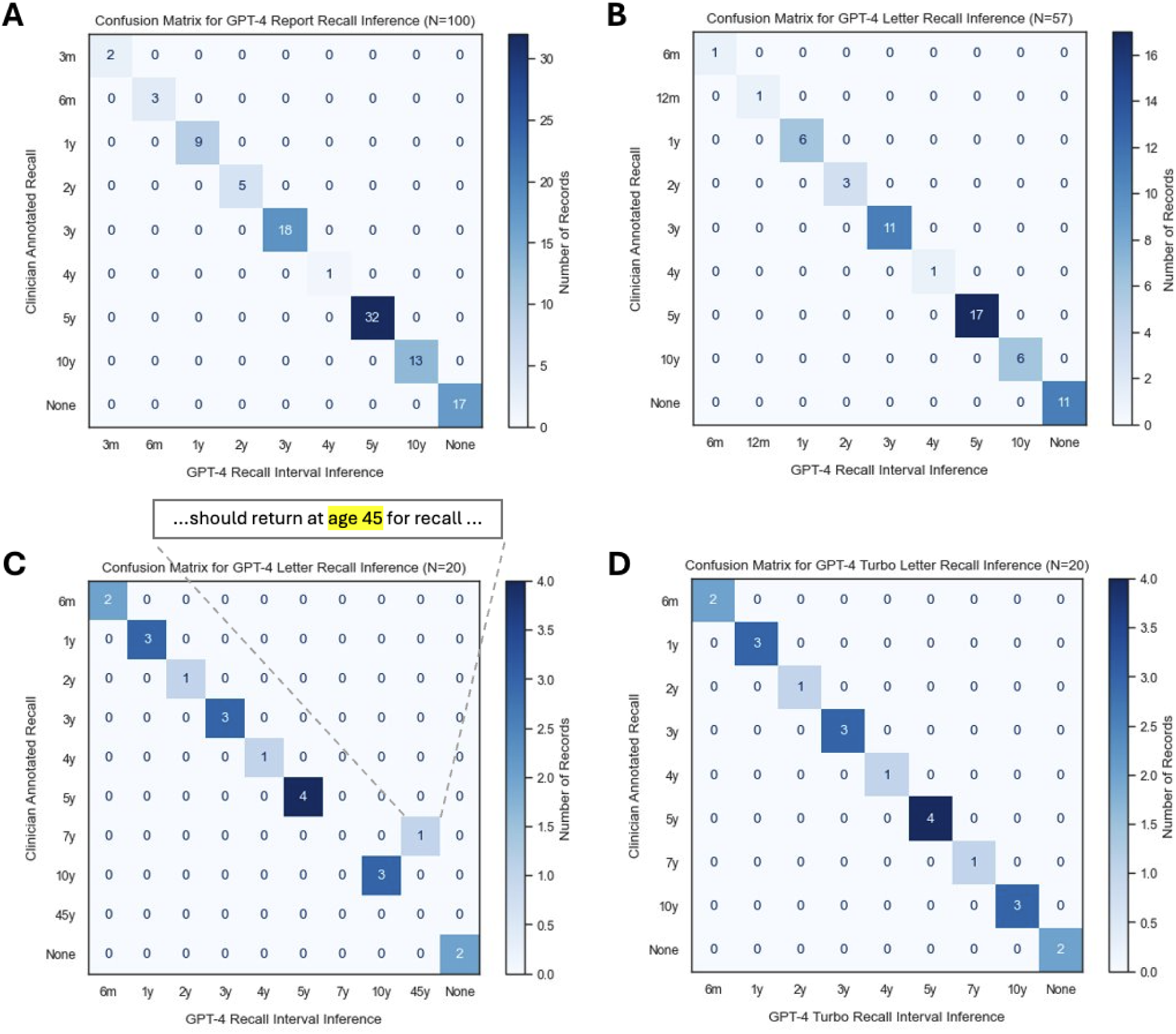
Confusion matrices for LLM GPT-4/GPT-4 Turbo inference performance compared to clinician review. Following optimization from smaller scale testing, results from formal assessment of performance of procedure reports (A) and patient letters (B) for N=100 patients showed perfect concordance with manual clinician assessment (Macro F1 = 1.0). C) Testing on a collection of 20 edge cases with dual colonoscopy and EGD recall demonstrated a failure mode of GPT-4 in which age at recall (45 years old) was misinterpreted as recall interval (45 years). D) Further prompt refinement and switching from GPT-4 to GPT-4 Turbo was able to rescue this failure while maintaining high performance on previously annotated data (confirming lack of performance regression). Original figure created by authors using data from hospital database (refer to methods section).Visualization designed using Adobe Illustrator.

To better explore performance on potential edge cases, we performed subsequent testing on a more challenging set of 20 cases from bidirectional endoscopies, as these require precise semantic distinction between the suggested recalls for the upper and lower endoscopic component. Although the model was able to correctly isolate the colonoscopy-relevant recall text, we did find a failure in which the model misinterpreted a recall age as a recall interval (Fig. 3C).

As one approach to more robustly handle this failure mode, we transitioned to GPT-4 Turbo, which demonstrated enhanced reasoning capabilities and successfully handled edge cases, including differentiating between target-age and interval-based recalls (Fig. 3D). Retrospective validation confirmed that GPT-4 Turbo maintained the high inference accuracy seen with GPT-4.

### Expanded Quality Control Including Anomaly Detection

As manual review becomes infeasible at larger scales, we implemented more efficient approaches for quality control at scale, namely field validation and anomaly detection. At intermediate scale (N=1000), field validation – which allows rapid examination of the unique values for an output field rather than all values – identified instances where invalid values (”PARSE ERROR”) were returned (often corresponding to patients who had aged out of surveillance), leading to further prompt revision (Fig. 2).

To detect even lower frequency errors at scale, we computed derived features (e.g., “age at recall,” “age at last colonoscopy”) and performed anomaly detection, in which we evaluated joint variable distributions for outliers (Fig. 4A). At full scale (N=118,061), we qualitatively identified outlier populations with unexpectedly short or long recalls, i.e., below 6 months or over 10 years, respectively. Small-value outliers typically corresponded to clinic follow-ups rather than colonoscopy follow-ups, while large values in which a patient was projected to return well beyond a standard screening age typically represented misinterpretation of absolute recall year as recall interval. These quality control methods identified rare failure modes: ∼1 in 1,000 (field validation) and ∼1 in 10,000 (anomaly detection).

**Figure 4.**
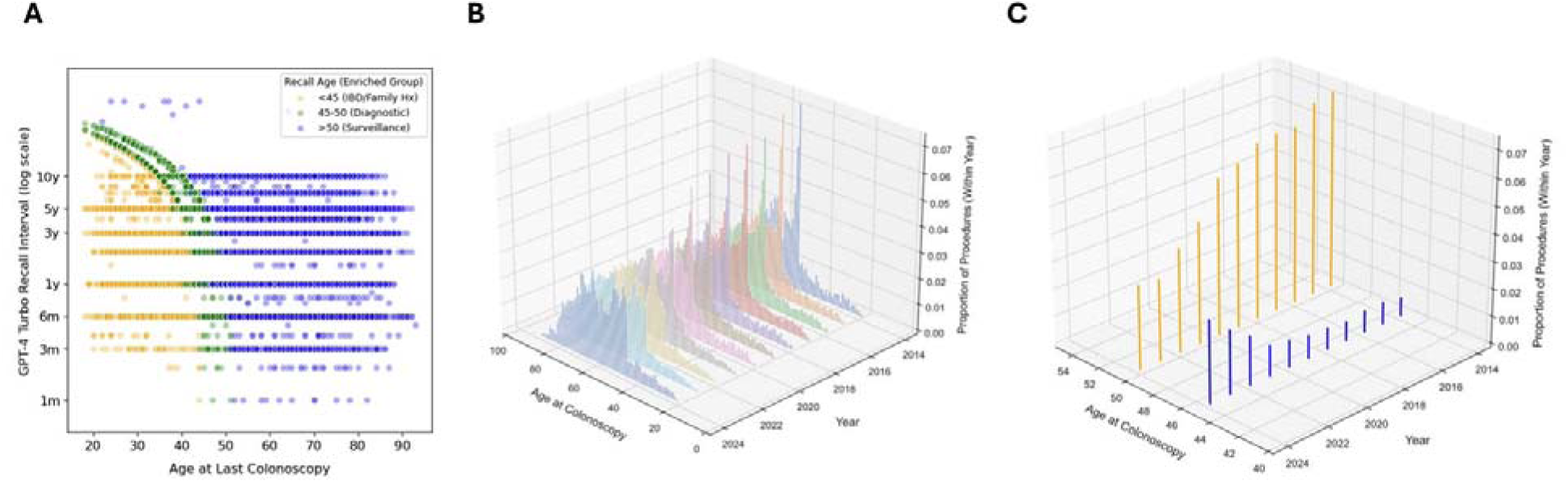
Exploratory data analysis of population-wide colonoscopy recall dataset. A) Comparison of calculated age at last colonoscopy and GPT-4 Turbo inferred recall interval. Points are colored by qualitative subgroup (yellow = next projected colonoscopy before age 45, including patients with IBD and family histories of CRC; green = patients projected to return for their next colonoscopy at a standard screening initiation age, enriched for diagnostic colonoscopies; blue = patients over age 50 with standard risk-based screening and surveillance intervals). B) Distribution of age at last colonoscopy over time in our institutional cohort. C) An increase in the proportion of patients being screened at age 45 coincides with the 2021 USPSTF guideline recommendation to move the age of screening initiation from 50 (orange) to 45 (blue).^5^ Original figure created by authors using data from hospital database (refer to methods section).Visualization designed using Adobe Illustrator.

### Real-World Deployment and Usability

Following implementation of quality control filters as above, structured AI-generated recall recommendations were uploaded into the new EHR’s health maintenance tab along with a brief automated comment citing the source type (report vs. letter) and date. This transparency enabled clinicians to quickly verify recall intervals and address discrepancies.

Post-deployment feedback from primary care physicians was collected by the IS team and was generally positive. One exception to this was a message inquiring about the absence of recall data for a patient whose procedure occurred at a low-volume affiliate—a known limitation of our pipeline anticipated to be quite uncommon across the cohort (<1% of procedures).

### Integrative Analysis at Scale

Given the unique opportunity to examine structured colonoscopy recall data for a massive clinical population, we explored what additional insights might be evident at scale. Further evaluation of groups in our anomaly detection plot identified three populations with differentially enriched groups (Fig. 4A):

1. Patients aged below 45 with recall dates projected to be before a standard screening start, representing those with family histories of colon cancer or IBD who often require more frequent surveillance.^4^
2. Patients with abbreviated recall intervals and next colonoscopy recommended between ages 45-50, representing those who had diagnostic colonoscopies with plans to return for standard age-based screening.
3. Patients with next colonoscopies at ages beyond this range were primarily those in standard risk-adjusted screening and surveillance pathways.

In a complementary analysis, we assessed the distribution of ages for patients undergoing colonoscopies for each year in the cohort (Fig. 4B). An increasingly prominent peak at age 45 could be appreciated relative to age 50 beginning in 2021 (Fig. 4C), corresponding to the USPSTF guideline change that year.^5^ These findings – reflecting both patient and provider level trends across time – hint at the potential insights made possible by harnessing LLMs within a broader end-to-end pipeline to structure data at scale.

## DISCUSSION

### Structured Data Transitions Create Critical Vulnerability Windows

A growing trend in healthcare involves the migration toward structured documentation systems—whether through individual health maintenance modules or full-scale EHR transitions. These changes aim to improve data fidelity and enable analytics, but they rarely come with automated mechanisms for transitioning legacy unstructured data. This results in a critical vulnerability window: the new fields may appear populated or reliable, yet important clinical information—such as prior surveillance recommendations—remains buried in unstructured notes. In this setting, clinicians may unknowingly omit essential tasks, assuming the structured data is accurate.

This case study highlights one such example: colonoscopy recall intervals, which were inconsistently populated in structured fields during a major EHR overhaul. Prior to intervention, over 70% of recall intervals would have had placeholder values that were larger than the clinically indicated interval—a gap that would have introduced risk of missed surveillance and potential diagnostic delays (Fig. 5). Our solution leveraged large language models (LLMs) to extract, validate, and restore these intervals, demonstrating how AI can mitigate transitional risk during EHR modernization efforts.

**Figure 5.**
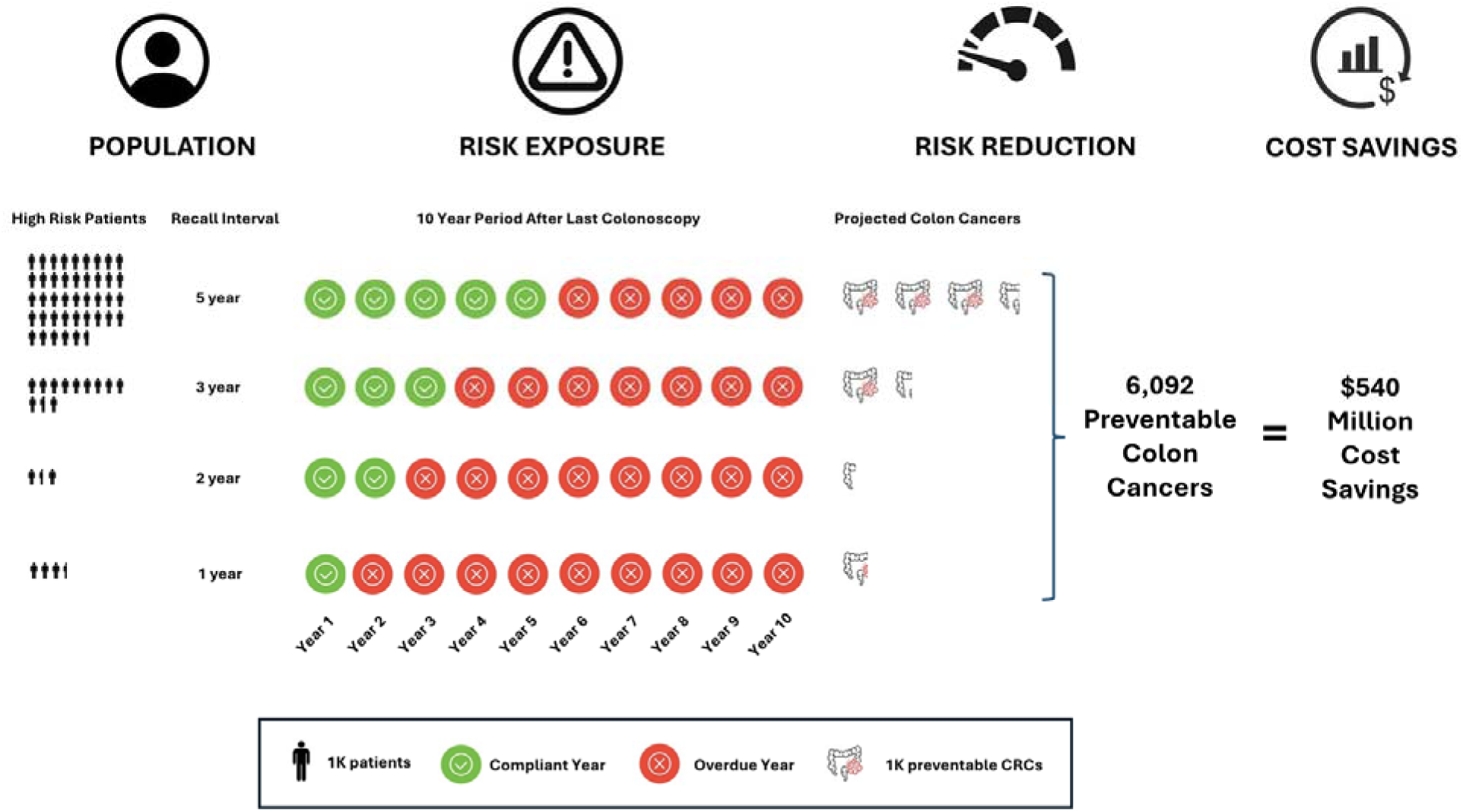
Potential impacts of an LLM-derived recall inference workflow on long term risk reduction and downstream cost savings relative to a non-individualized 10-year recommendation. Patients were considered high-risk if their individualized recall recommendation from their last colonoscopy was 5 years or less (so less than e.g., a standard 10-year recommendation). Each year of the following decade after the last colonoscopy is annotated based on whether a patient assigned to a default 10-year interval would be compliant with the last recommendation or not. Projected colon cancers for each risk group are displayed (using a baseline 2% / year rate of CRC in overdue patients and assuming an efficacy of 75% for colonoscopy).^6,7^ An average cost of CRC management of $80,000/patient was used to model total cost savings.^12^ Original figure created by authors using data from hospital database (refer to methods sections), with parameters adapted from Sullivan et al. (2022), Chen et al. (2003), and Yabroff et al. (2008). The figure was created using Adobe Illustrator.

### LLMs as Transitional Infrastructure in the Era of Structured Data

Our pipeline was a multi-layered system designed to ensure fidelity and scalability (Fig. 1). Although deployed in a batch mode during our institution’s EHR transition, the pipeline was built with transparent traceability in mind. Clinicians were able to view AI-extracted recall intervals alongside supporting text from the original reports, enabling real-time verification and clinical trust. This human-in-the-loop design supports not just migration, but ongoing quality assurance.

Unlike traditional NLP efforts focused on classification or keyword extraction, our approach targeted higher-order reasoning across diverse documentation styles. It reconciled conflicting data, enabled structured field completion at scale, and did so with sufficient clarity to allow for human validation. This suggests a generalizable paradigm for other areas of healthcare—such as mammography, bone density screening, or even vaccine catch-up documentation—where historical free-text data must be ported reliably into structured formats.

### Cost Avoidance and System-Level Impact

The financial implications are significant. By enabling accurate patient-specific recalls rather than default 10-year intervals, we estimate preventing up to 6,092 colorectal cancer cases, representing 406,160 additional patient-years of surveillance (Fig. 5).^6,7^ These figures translate to substantial morbidity reduction as well as a projected cost savings between $400-670 million (Fig. 5), a substantial return on investment considering inference and OCR costs totaling under $20,000 for one run of the pipeline.^8^ There may also be associated improvements in preventative health equity given that at-risk populations might be most vulnerable to loss to follow up in the setting of inappropriately long surveillance intervals.^9^

### End-to-End Workflow Integration

A key strength of our system is its design as an end-to-end solution for structured data backfill. While the current implementation operates as a batch pipeline—leveraging a large language model with Python-based post-processing to support retrospective migration during the EHR transition—it is embedded within a broader clinical workflow. The output is integrated into physician-facing interfaces, allowing providers to trace each structured recall recommendation back to its original source note for validation. This design ensures transparency, supports clinical oversight, and lays the foundation for future real-time or continuous NLP integration. Unlike prior NLP efforts focused solely on classification, our approach enables scalable chart abstraction with human-in-the-loop validation, making it well suited for multi-institutional deployment and continuous quality improvement.

### Limitations

Our study reflects a single academic medical center’s experience, potentially limiting generalizability. Our legacy EHR (webOMR) was one of the earliest systems developed, so pre-processing challenges might differ in alternative systems.^10^ We relied primarily on GPT-4/GPT-4 Turbo; newer reasoning enabled models might improve accuracy but increase costs.^11^ Finally, while we executed two batch processes at scale, we did not deploy a prospective recall system, which could surface additional implementation challenges (e.g., real-time access, fault tolerance redundancy, and proactive patching).

### Conclusion

The migration to structured data fields in clinical documentation represents a double-edged sword, with the potential for long-term improvements in patient outcomes at the cost of potentially increased risk in the short term. Our LLM-driven colonoscopy recall system demonstrates how modern tools can bridge this gap when deployed within broader end-to-end pipelines and underscores the potential of transformer-based architectures to bring unprecedented levels of precision to diverse aspects of clinical care.

## AUTHOR CONTRIBUTIONS

A.M., R.P., S. W., H.H, J.S., A.S., T.F., D.R, A.M., V.J, A.R., J.D.F. participated in primary data analysis, pipeline development, and interpretation. G.P. and A.S. participated in generation of structured views of legacy data. A.M., R.P., M.J.A., S.F., A.R., J.D.F. participated in review of clinical data. A.M., V.J., A.R., and J.D.F participated in project supervision. A.M., R.P., S.W., A.R., and J.D.F. participated in primary drafting of the manuscript. All authors participated in the final assembly and revision of the manuscript.

## Data Availability

All data produced in the present study are available upon reasonable request to the authors.
All data produced in the present work are contained in the manuscript.

## ACKNOWLEDGEMENTS

The authors would like to thank the many patients whose data is reflected in this study, and without whom this work would not be possible. In addition, we would like to thank colleagues from the Beth Israel Deaconess Medical Center (BIDMC) Administration, Information Systems team, and the Gastroenterology Division, for helpful feedback in designing and implementing this project. Insights regarding the integration of AI in clinical workflows was also appreciated from the BILH AI Innovation Task Force.

## CONFLICT OF INTEREST STATEMENT

A.R. is the founder of Halo Solutions, LLC, which provides consulting services related to applications of artificial intelligence for healthcare data. The remaining authors declare no competing interests related to this study.

## Funding

This study was supported internally using institutional funds at Beth Israel Lahey Health (BILH).

## Affiliations

**Aman Mohapatra, Rachel Porth, Maelys J. Amat, Sarah Flier, David Rubins, Amy Miller, and Joseph D. Feuerstein**

*Department of Medicine, Beth Israel Deaconess Medical Center, Harvard Medical School, Boston, MA, USA*

**Si Wong, Gail Piatkowski, John Shang, Adam Salsman, Ted Fitzgerald, Ayad Shammout, David Rubins, Amy Miller, and Venkat Jegadeesan**

*Information Systems, Beth Israel Lahey Health, Boston, MA, USA*

**Heather Hardway and Arvind Ravi**

*Halo Solutions, LLC, USA*

## Notes

### Competing Interest Statement

The authors have declared no competing interest.

### Author Declarations

The Committee on Clinical Investigations of Beth Israel Deaconess Medical Center gave ethical approval for this work.

## REFERENCES

1. Sedlakova J, Daniore P, Horn Wintsch A, et al. Challenges and best practices for digital unstructured data enrichment in health research: A systematic narrative review. PLOS Digit Health. 2023;2(10):e0000347. doi:10.1371/journal.pdig.0000347

2. Maleki Varnosfaderani S, Forouzanfar M. The Role of AI in Hospitals and Clinics: Transforming Healthcare in the 21st Century. Bioeng Basel Switz. 2024;11(4):337. doi:10.3390/bioengineering11040337

3. Reisman M. EHRs: The Challenge of Making Electronic Data Usable and Interoperable. P T Peer-Rev J Formul Manag. 2017;42(9):572–575.

4. Clarke WT, Feuerstein JD. Colorectal cancer surveillance in inflammatory bowel disease: Practice guidelines and recent developments. World J Gastroenterol. 2019;25(30):4148–4157. doi:10.3748/wjg.v25.i30.4148

5. US Preventive Services Task Force, Davidson KW, Barry MJ, et al. Screening for Colorectal Cancer: US Preventive Services Task Force Recommendation Statement. JAMA. 2021;325(19):1965-1977. doi:10.1001/jama.2021.6238

6. Sullivan BA, Noujaim M, Roper J. Cause, Epidemiology, and Histology of Polyps and Pathways to Colorectal Cancer. Gastrointest Endosc Clin N Am. 2022;32(2):177–194. doi:10.1016/j.giec.2021.12.001

7. Chen CD, Yen MF, Wang WM, Wong JM, Chen THH. A case-cohort study for the disease natural history of adenoma-carcinoma and de novo carcinoma and surveillance of colon and rectum after polypectomy: implication for efficacy of colonoscopy. Br J Cancer. 2003;88(12):1866–1873. doi:10.1038/sj.bjc.6601007

8. Yabroff KR, Mariotto AB, Feuer E, Brown ML. Projections of the costs associated with colorectal cancer care in the United States, 2000-2020. Health Econ. 2008;17(8):947-959. doi:10.1002/hec.1307

9. Agunwamba AA, Zhu X, Sauver JSt, Thompson G, Helmueller L, Finney Rutten LJ. Barriers and facilitators of colorectal cancer screening using the 5As framework: A systematic review of US studies. Prev Med Rep. 2023;35:102353. doi:10.1016/j.pmedr.2023.102353

10. Sands DZ, Rind DM, Safran C. Dissemination of electronic patient records using primary care referrals as a vector for change. Stud Health Technol Inform. 2001;84(Pt 1):685–689.

11. Klang E, Apakama D, Abbott EE, et al. A strategy for cost-effective large language model use at health system-scale. NPJ Digit Med. 2024;7(1):320. doi:10.1038/s41746-024-01315-1

